# *CYP2B6*\**6* single nucleotide polymorphism among patients with uncomplicated malaria in Adjumani district, Uganda: implications on efficacy of artemether-lumefantrine

**DOI:** 10.1101/2025.06.13.25329585

**Authors:** Martin Kamilo Angwe, Norah Mwebaza, Sam Lubwama Nsobya, Ronald Kiguba, Patrick Vudriko, Savior Dralabu, Denis Omali, Maria Agnes Tumwebaze, Moses Ocan

## Abstract

CYP2B6, one of the most polymorphic enzymes, plays a major role in the metabolism of artemisinin and its derivatives. Variation in the frequency of the most common yet functionally deficient *CYP2B6*6* allele may impact artemisinin exposure, potentially contributing to differences in treatment outcomes. This study assessed the prevalence of the *CYP2B6*6* genotype among patients with uncomplicated malaria treated with artemether-lumefantrine at Adjumani District Hospital in the West Nile region, Uganda.

A total of 100 randomly selected patients with microscopically confirmed uncomplicated *P. falciparum* malaria receiving artemether-lumefantrine (AL) were included in the study. Blood samples, 2-3 mL each, were collected using EDTA tubes on days 0 and 3 after AL administration. DNA was extracted using Qiagen DNA Mini kit. Polymerase Chain Reaction-Restriction Fragment Length Polymorphism (PCR-RFLP) was used to determine the *CYP2B6*6* genotype of the participants. *P. falciparum* positivity was determined by microscopy and qPCR. For qPCR, parasite clearance was determined using comparative CT value. Data analysis was done using STATA *ver* 17.0 at a 95% significance level.

Among the malaria patients genotyped, 65% were female, and 35% were male, with a mean age of 17±10 years. The *CYP2B6*6* variant allele frequency was 0.37, and the genotype frequency was 43% GG, 17% TT and 40% GT. *P. falciparum* day 3 microscopy positivity was 14% (6/43) among the GG, 37.5% (15/40) among the GT, and 17.64% (3/17) among the TT patients. PCR positivity rates were 58.1% (25/43) in the GG, 72.5% (29/40) in the GT, and 52.9% (9/17) in the TT patients. Heterozygous individuals (GT) had slow parasite clearance by a 10-fold difference compared to the homozygous (GG, TT) individuals (*U*=323.0; *Z*=-2.3442; *p*=0.019).

Malaria patients in Adjumani district had a high frequency of the *CYP2B6*6* variant allele. Interindividual variability in *P. falciparum* clearance exists, with heterozygous patients demonstrating slow artemether-lumefantrine *P. falciparum* parasite clearance. There is a need to consider incorporating host genomic factors, such as metabolising enzyme genotype into routine Therapeutic Efficacy Studies (TES).

## Introduction

Malaria is one of the most devastating infectious diseases globally, with an estimated case of 263 million in 2023, showing an increased risk and cases from the previous year. *P. falciparum* accounts for at least 95% of malaria-related cases and deaths in sub-Saharan Africa (1). Although the 2024 malaria report estimated an increase of 11 million in malaria cases (1), there has been a decrease in malaria-associated mortality owing to the availability of diagnostic services and prompt treatment with antimalarial drugs. Uganda accounts for at least 5% of the global malaria cases. It remains among the top 5 malaria-endemic countries despite intensive control interventions such as Indoor residual spraying, insecticide-treated mosquito nets and the availability of Artemisinin-based combination therapy for case management.

Artemisinin-based Combination Therapies (ACTs) are the current cornerstone of malaria treatment globally. The World Health Organisation recommends artemisinin-based combination therapy and IV artemisinin agent as first-line therapies for uncomplicated and severe malaria, respectively (1–3). Artemisinin and its derivatives have a short half-life; however, their activity is complemented in various combinations by different long-acting partner drugs such as lumefantrine, piperaquine, and amodiaquine (4,5). Uganda adopted AL as the primary first-line therapy and dihydroartemisinin-piperaquine as a second-line. Recent reports from Uganda indicate treatment failures with AL and *K13* mutations associated with slow parasite clearance (6–9). Artemether is the artemisinin derivative with a plasma half-life of 1-3 hours and is responsible for the rapid reduction in malaria parasite biomass, while the partner drug, lumefantrine, with a longer half-life of 3-4 days, are responsible for the clearance of residual mass in *Plasmodium falciparum* parasites (10–12).

Artemether is metabolised by Cytochrome P450 (CYP) enzymes to its active and potent metabolite, dihydroartemisinin (DHA), in phase 1 reaction (13–16). A study by Honda *et al.* demonstrated that CYP2B6*6 is the primary enzyme responsible for artemether demethylation to DHA (17). DHA is further metabolised by Uridine 5’-diphospho-glucuronosyltransferase (UGT) in the phase 2 reaction, particularly UGT1A9 and UGT2B7 (18,19). The contribution of polymorphic CYP2B6 in artemether demethylation may pose significant individual variability in artemether conversion to DHA (20–23). The *CYP2B6*6* alleles and the resulting varying DHA levels may impact parasite clearance during malarial treatment in some subgroups due to the differences in CYP2B6 activity. The polymorphic CYP2B6 enzyme is encoded by the *CYP2B6* gene located on chromosome 19 (24,25) and contributes to the phase one metabolism of antimalaria drug (artemisinin) and several other drugs (25).

Single Nucleotide Polymorphisms (SNPs) in *CYP2B6*6* are associated with altered Artemisinin drug metabolism and clearance rates (26–28). The *CYP2B6*6* SNP, characterised by a G-to-T substitution at position 516, reduces enzymatic activity. This potentially leads to slower drug activation of artemether, which could affect *P. falciparum* parasite clearance. According to Hofmann, 516G>T causes irregular splicing, decreasing expression and activity in CYP2B6*6 (29). The polymorphic CYP can be categorised as fast, intermediate, and slow metabolisers. The fast metabolisers are the GG genotypes (wild type), while the intermediate and slow metabolisers are the 516G>T, i.e., GT and TT, respectively (20). The African population largely has *CYP2B6c516G>T* (rs3745274), with at least 50% of some ethnic groups being intermediate and slow metabolisers of CYP2B6 substrates (30–33). The high prevalence of the *CYP2B6*6* allele means that artemether may not achieve optimal malaria parasite clearance. There is inadequate information on the implication of *CYP2B6*6* variability on the efficacy of AL in malaria treatment in endemic settings.

In Uganda, recent studies have reported the prevalence of slow artemisinin-clearing *P. falciparum* parasites in northern Uganda (6,7). The efficacy of artemisinin agents is routinely studied and monitored; however, host genomics is not included in most studies. This study evaluated the distribution of *CYP2B6*6* genotypes and the potential correlation with *P. falciparum* parasite clearance among malaria patients treated using AL at Adjumani District Hospital, Uganda. Understanding how the host’s genetics, particularly *CYP2B6*6* polymorphisms, influence parasite clearance provides insight into interindividual variability in ACT treatment. This could help inform the optimisation of ACT dosing regimens in diverse malaria-endemic populations.

## Materials and Methods

### Ethical statement

Ethical approval was obtained from the School of Biomedical Science Research and Ethics Committee, College of Health Science, Makerere University (SBS-2022-157). Administrative clearance was obtained from the district health officer to conduct the study in the hospital. Informed consent (18 years old and above), parental consent (5-7 years old), and assent plus parental consent (8-17 years old) were obtained.

### Study design and setting

This cross-sectional study was nested in a prospective study described in our prior publication among patients aged five years and above who were prescribed standard artemether-lumefantrine to treat uncomplicated *P. falciparum* malaria at Adjumani District Hospital (6). Adjumani district is a predominantly Nilo-Hamites population with a high prevalence of *P. falciparum* malaria, with year-round infections (34). The study was conducted between 09/September/2022 and 06/November/2022. A total of 109 patients consented to participate; 7 did not turn up on day 3 for the last sampling, and 2 reported a change of treatment; hence, 100 patients were included in the study. Venous blood (2-3 mL) was collected in EDTA tubes on day 0 before AL treatment and on day 3 after treatment. Thick and thin blood smears were prepared on the spot for microscopy. Laboratory analysis was performed at the College of Veterinary Medicine, Animal Resources and Biosecurity, the genomic lab of the Pharmacology/Research Centre for Tropical Diseases and Vector Control (RTC), and Central Diagnostic Laboratories (CDL).

### DNA extraction and processing

DNA was extracted from 200 µl heparinised blood samples using a Qiagen DNA mini kit (QIAamp DNA Blood Mini Kit-250, Germany) according to the manufacturer’s guidelines. The DNA extract was eluted in 50 μL elution buffer, and a spectrophotometer (NanoDrop Lite Plus, Thermo Scientific) was used to quantify the DNA (25–40 ng/μl) before the downstream procedure of CYP2B6*6 genotyping and *P. falciparum* qPCR.

### CYP2B6*6 Genotyping

To determine the allele frequency of *CYP2B*6* alleles, PCR-RFLP was performed using patient DNA samples. Reaction mixture of 25 µl was prepared comprising of 12.5 µl OneTaq 2X Master Mix (NEB, New England BioLabs Inc), 1 µl of 10 (µM) each forward and reverse primer (35) (*CYP2B6-F:* 5′-TCTCGGTCTGCCCATCTATAAACT-3′ and *CYP2B6-R*: 5′-CCTGACCTGGCCGAATACA-3′), 9.5-9.88 µl of nuclease-free water and 2.5 ng of the DNA template. Amplification conditions comprised 1 cycle of 95 ^°^C initial denaturation for 5 minutes, followed by 30 cycles of 95^°^C denaturation for 30 seconds, 56 ^°^C annealing for 30 seconds, 65 ^°^C extensions for 60 seconds and 1 cycle of final extension at 65 ^°^C for 5 minutes. The PCR product was digested using the BSR1 enzyme in a reaction mixture containing 0.3 µl of enzyme, 1.5 µl of buffer, 1.2 µl of PCR water, and 12 µl of the DNA amplicon. The reaction was then incubated at 65 ^°^C overnight (∼13 hours). The reaction was stopped by heating at 80 ^°^C for 15 minutes. Electrophoresis was performed at 100 volts for 30 minutes with 2% agarose gel; DNA was stained with 0.05 µg/mL Acridine orange (SafeView Classic) and visualised using Utra Violent light (GEL Doc Biobase).

### Parasite clearance determination

Parasitemia was determined by microscopy and qPCR, as reported in our previous study (6). Microscopy positivity on days 0 and 3 was determined, and qPCR was used to determine the parasite clearance using comparative CT values through fold change as previously described (6,36). Briefly, day 0 and day 3 microscopy was performed by staining the thick and thin smears with 10% Giemsa stain for 30 minutes and examined by a light microscope (Olympus) under 100X magnification by two independent laboratory technologists.

A real-time PCR system (QuantStudioTM5) was used to perform the qPCR using previously described primers and probes based on the 18S rRNA (37) sequence manufactured by Inqaba Biotec Laboratory, South Africa. The specific primers used were 5’-AGCAGGTTAAGATCTCGTTCG-3′ as the forward and 5′-GCTCTTTCTTGATTTCTTGGATG-3 as the reverse primer; the probe was 5′-ATGGCCGTTTTTAGTTCGTG-3′ labelled with 5′FAM (6-carboxyfluorescein) as the reporter and BHQ-1TM (Black Hole Quencher) as the quencher. The qPCR master mix (Luna Universal Probe qPCR Master mix NEB, New England BioLabs Inc) was used to prepare 20 µl reaction mixture, accordingly, comprising 0.8 µl each primer, 0.4 µl of the probe, 20 ng of the template DNA and variable nuclease-free water to make 20 µl. Amplification was performed with initial denaturation at 94 ^°^C for 5 minutes, 40 cycles of 30 seconds denaturation at 94 ^°^C, annealing at 54 ^°^C for 90 seconds, and extension for 90 seconds at 68^°^C, followed by final extension for 90 seconds at 68 ^°^C. PCR was performed in duplicate, and CT values were recorded and used to determine the parasite clearance through fold change in patients who remained PCR positive for *P. falciparum* on day 3.

### Data management and analysis

Genotype and allele data were analysed and presented as counts, frequencies and percentages using STATA *ver* 17. Allele and genotype distribution in the population was determined by chi-square test at a 95% confidence interval; the power of association was measured through the phi coefficient and Chalmer’s V. A non-parametric test was used to compare parasite clearance among the different genotypes using a log-transformed fold reduction using Mann-Whitney U test with a 95% level of significance. Parasite clearance is presented as fold reduction in parasitemia between day 0 and day 3, determined using comparative CT value, 2^−ΔΔCT^, on day 3 *P. falciparum* PCR positives. The fold reduction and parasitemia values are presented as median and interquartile range.

## RESULTS

### Characteristics of study participants

A total of 100 malaria patients were included in the study. The patients’ characteristics were reported in our previous study (6). Briefly, the study population comprised 65% females (65/100) and 35% males (35/100) aged 5 to 46 years, with a mean of 17±10 years. The initial parasitemia was 3868, 11104 (Geometric Mean, IQR) by microscopy before treatment (day 0).

### CYP2B6*6 distribution among the malaria patients

The amplification of the *CYP2B6*6* gene produced a band size of 400bp before digestion. Upon digestion with the BSR1 enzyme, three bands were produced for the wild type, two for the heterozygous genotypes and one for the homozygous variant genotype (Figure 1). The allele frequency of *CYP2B6*6* was 0.37 for the T (minor) alleles, and the genotype frequency for *CYP2B6*6* was 43% GG, 17% TT and 40% GT among the study participants. The minor allele frequency is higher in male (0.49) patients than in females (0.31) (Table 1). The frequencies of *CYP2B6*6* genotypes in the malaria patients aligned with Hardy-Weinberg equilibrium (*X*^2^=0.9954, *p*=0.6) (Table 2).

**Figure 1:**
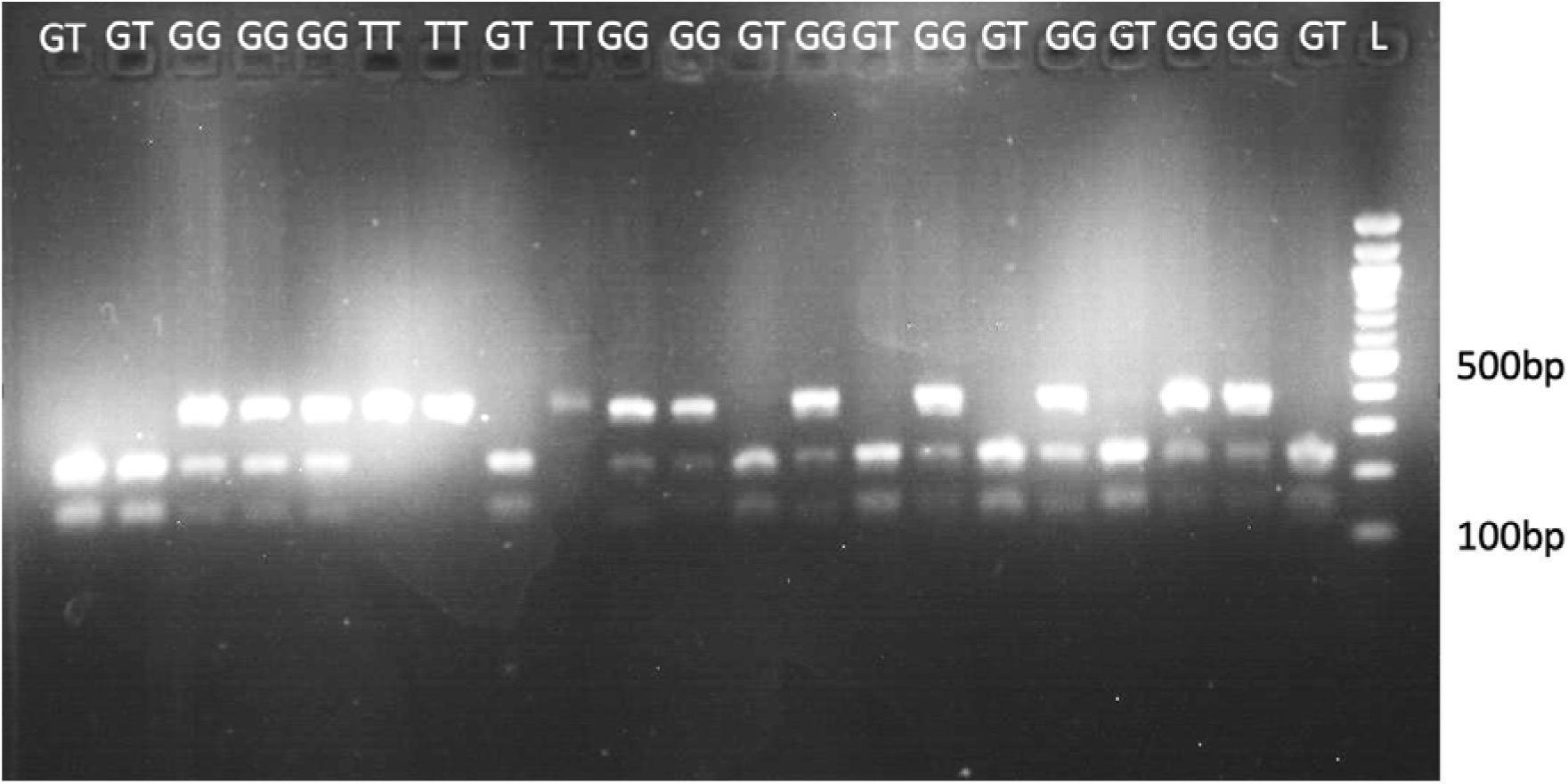
Gel image of *CYP2B6*6* after digestion with BSR1: GG=homozygous wild-type, GT=heterozygous variant, TT=homozygous variant, L=100bp DNA ladder.

**Table 1:** Distribution of alleles and genotype among malaria patients.

| Sex | Allele |  |  |  |  | Genotype |  |  |  |  |  |
| --- | --- | --- | --- | --- | --- | --- | --- | --- | --- | --- | --- |
| | G | T | $X^2$ | *P-value* | $\phi$ | GG | GT | TT | $X^2$ | *P-value* | $\phi_c$ |
| Female | 0.69 | 0.31 | 6.0208 | 0.0141 | 0.1735 | 0.47 | 0.43 | 0.1 | 12.3327 | 0.0021 | 0.2483 |
| Male | 0.51 | 0.49 |  |  |  | 0.26 | 0.5 | 0.24 |  |  |  |
| **Frequency** | **0.63** | **0.37** |  |  |  | **0.43** | **0.4** | **0.17** |  |  |  |

**Table 2:** Observed and expected genotype frequency in malaria patients.

| Genotype | Observed | Expected | $X^2$ | *P-value* |
| --- | --- | --- | --- | --- |
| GG | 43 | 40 | 0.9954 | 0.6 |
| GT | 40 | 46 |  |  |
| TT | 17 | 14 |  |  |

### Association of the *CYP2B6*6* allele with parasite clearance after 3 days of AL treatment

The initial parasitemia (day 0) among the patients stratified by genotypes (GG, GT, and TT) (Geometric mean, IQR) was 2319, 20323 for GG, 1807, 8188 for GT and 7613, 10960 for the TT patients. The day 3 microscopy positivity was 14% (6/43) among the GG, 37.5% (15/40) among GT and 17.64% (3/17) among TT patients. PCR positivity was 58.1% (25/43) in the GG, 72.5% (29/40) in the GT and 52.9% (9/17) in the TT *CYP2B6*6* patients.

Parasite clearance differed among the *CYP2B6* genotypes (Table 3). The fold reduction (median, IQR) was 1020, 15504 in GG patients, 1941, 8868 in the TT patients and 105, 2354 in the GT patients among the patients who remained PCR positive for *P. falciparum* on day 3. Parasite clearance was lowest among the GT patients, with 10-fold lower parasite clearance (fold change) than the GG and TT patients. Interindividual variability in parasite clearance was observed among the malaria patients; heterozygous individuals had slow parasite clearance, GG Vs GT genotype (p=0.036) and GG+TT Vs GT genotype (*p*=0.019).

**Table 3.** Day 3 *P. falciparum* among the different genotypes and parasite clearance.

| Genotypes and the combination | Day 3 microscopy positive<br>N=100 |  |  | Day 3 qPCR positive<br>N=100 |  |  | Parasite clearance/ CT value<br>Fold change n=63 |  |  |
| --- | --- | --- | --- | --- | --- | --- | --- | --- | --- |
|  | OR | CI (95%) | *P-value* | OR | CI (95%) | *P-value* | *U* | *Z* | *P-value* |
| GT, GG | 3.70 | 1.26, 10.63 | 0.02 | 0.53 | 0.21, 1.32 | 0.25 | 241.0 | -2.1077 | 0.036 |
| TT, GG | 1.32 | 0.29, 6.02 | 0.7 | 0.81 | 0.26, 2.50 | 0.78 | 111.0 | -0.0586 | 0.97 |
| (GT+TT),<br>GG | 2.02 | 1.02, 7.95 | 0.058 | 1.44 | 0.64, 3.27 | 0.41 | 352.0 | -1.7280 | 0.09 |
| GT,<br>(GG+TT) | 3.40 | 1.31, 8.83 | 0.016 | 2.02 | 0.85, 4.77 | 0.14 | 323.0 | -2.3442 | 0.02 |

OR=odds ratio (crude), CI=confidence interval at 95%, n for the subjects is GG=25, GT=29,
TT=9, N=Number of subjects, n=number of subjects who remained day 3 qPCR positive for
*P. falciparum*.

## Discussion

This study investigated the distribution of the *CYP2B6*6* allele and *P. falciparum* clearance in symptomatic uncomplicated malaria patients prescribed artemether-lumefantrine. The frequency of the *CYP2B6*6* variant allele was 0.37 (37%), and the patients’ genotype frequency was 43% (GG), 40% (GT), and 17% (TT). There was a high interindividual difference in day 3 parasitemia (microscopy) and parasite fold reduction (*P. falciparum* clearance); patients with the GT genotype had 10-fold lower parasite clearance than those with the GG or TT genotype.

The *CYP2B6*6* allele frequency of the African population, in general, is around 65% for the G allele and 35% for the T allele (38,39), which is comparable to our finding that reported a 37% frequency of the T allele. A study in Mozambique demonstrated significant variation in the distribution of various *CYP2B6* variants, especially the G516T substitution (40). Similarly, studies in Uganda and Zimbabwe reported a high frequency of the *CYP2B6*6* variant among the African population (41,42). The frequency of the *CYP2B6*6* allele (0.37) in our studied population was relatively similar to those reported among the Chinese population but generally higher than those observed in Caucasians (∼0.256) and Japanese (∼0.164) (43–45). The high frequency of the variant allele is a concern to AL therapy as the CYP2B6 enzyme is key in the metabolism of artemether to DHA. The low expression of this enzyme means individuals with such a variant allele will have subtherapeutic DHA levels, which could affect malaria parasite clearance.

We demonstrated a significant interindividual variation in parasite clearance, with *CYP2B6*6* heterozygous (GT) patients showing 10-fold lower P. falciparum clearance than the wild-type (GG). The high frequency of the *CYP2B6\**6 variant allele and its association with slow parasite clearance among GT patients suggest that many Ugandan populations are intermediate/poor metabolisers of artemether. This could lead to subtherapeutic levels of DHA, resulting in poor malaria treatment outcomes, as observed among the patients with heterozygous (GT) genotypes. The relationship between the patient *CYP2B6*6* genotype and parasite clearance in malaria patients has not been documented to highlight the effect of the genotype on parasite clearance. However, evidence from *in vitro* studies points to *CYP2B6*6* as a critical determinant of artemether metabolism to the most active metabolite, DHA (17,46). Previous studies demonstrated poor treatment outcomes, although attributed to the pharmacokinetic differences in lumefantrine (47,48). Comparative studies on the efficacy of AL and DHA-Piperaquine demonstrate slight superiority in DHA-PQ over AL therapy (49–51). Notably, Uganda’s Ministry of Health recommends that DHA and its partner drug be used as a second-line therapy for certain individuals when AL fails (51).

All patient groups had significant illness resolution, even though the parasite clearance varied amongst the malaria patients; some even had 0% parasitemia on day 3 despite having the variant *CYP2B6*6* genotype. However, the observed clearance across the patients might be explained by the combined efficacy of artemether and the companion medication lumefantrine. Artemisinin combination therapy has generally remained efficacious in Uganda, with no known resistance reported for the partner drug, lumefantrine (51,52). The partner drug may thus mask the reduced efficacy of the artemether component of the ACT. Meanwhile, some patients demonstrated no parasite clearance on day 3 across all the genotypes; a study in the region demonstrated the emergence of partial resistance against artemisinin (6,7,53). The combined effect of *CYP* and *UGT* alleles can equally affect parasite clearance; UGT is the key enzyme in the Phase 2 metabolism of DHA (19). However, polymorphisms of functional significance also occur in *UGT1A9* and *UGT2B7* genes in Africa (18), whose products are involved in DHA pharmacokinetics and pharmacodynamics. Other non-genetic factors could also affect AL parasite clearance, such as food intake, disease status, drug-drug interaction and sex (4,16,54,55).

There were some limitations associated with this study. Firstly, pharmacokinetic parameters were not taken to quantify artemether or its metabolite DHA in the blood. Secondly, the study only considered CYP2B6*6, but other enzymes such as UGT and other CYPs may play a part in determining the therapeutic effect of artemether. Also, the small sample size of the patients with the TT genotype may mask the actual value for this category. However, we believe the sample size was adequate upon which a baseline conclusion could be drawn; although pharmacokinetic parameters were not determined, this could not affect the study outcome as the results were based on the parasite clearance from CT values, which were paired (day 0 and day 3).

## Conclusion

High *CYP2B6*6* interindividual variability is evident in malaria patients in Adjumani district. Heterozygous patients demonstrated delayed parasite clearance after treatment with AL. Host genetic variability could determine malaria treatment outcomes with artemether-lumefantrine. Thus, there is a need to explore the effect of host genetic variability in artemether therapy during efficacy studies and to understand the drivers of persistent *P. falciparum* after treatment with AL and the development of artemisinin resistance. Future studies could stratify large sample sizes by genotype, consider other artemisinin-metabolising enzymes, and pharmacokinetic parameters.

## Data Availability

All data produced in the present study are available upon reasonable request to the authors

## Acknowledgement

We would like to thank the research participants from Adjumani Hospital, the staff of Adjumani Hospital and colleagues at the Pharmacology/Research Center for Tropical Disease and Vector Control (RTC) laboratory and Central Diagnostic Laboratory.

## Authors’ contributions

The authors, MKA, MO, NM and SLN conceptualised the study. Field data collection was conducted by MKA. Laboratory analysis was done by MKA, SD, and MAT. The analysis of the data was done by MKA, MO and NM. The first draft of the manuscript was written by MKA. Review of the draft manuscript was done by MO, NM, VP, RK, DO, SD and MAT. All authors read and approved the final version of the manuscript.

